# Bridging the Health Literacy Gap: Women and Community Stakeholders’ Perspectives on Maternal and Child Health Communication in Peri-urban Karachi, Pakistan

**DOI:** 10.1101/2025.09.02.25334971

**Authors:** Sameeta Kumari, Ayesha Khalid, Hajra Malik, Mahim Maher, Muhammad Imran Nisar, Fyezah Jehan, Zahra Hoodbhoy

**Affiliations:** Department of Paediatrics and Child Health, Aga Khan University Hospital, Karachi, Pakistan

**Keywords:** Health literacy, Health message dissemination, Community health workers, Peri-urban communities

## Abstract

Health literacy significantly influences health outcomes, particularly in Low-and Middle-Income Countries. Communication of maternal and child heath messages occurs via text messages, phone calls, television, newspaper articles, and home visits. An exploratory qualitative study was conducted in four peri-urban sites of Bin Qasim Town, Karachi to explore the perspectives of women and community stakeholders on effective health messaging strategies in peri-urban Karachi, Pakistan. Seven focus group discussions (FGDs) with women of reproductive age, and seven key informant interviews (KIIs) with the community stakeholders were conducted. Women were randomly selected from the surveillance database, while stakeholders were purposively sampled. Audio-recorded FGDs and KIIs were manually transcribed and analysed using thematic analysis. Three major themes emerged: (i) the growing but constrained role of digital platforms, (ii) facilitators and barriers of health message dissemination, and (iii) community-suggested solutions. While digital tools like WhatsApp and TV were popular among smartphone users, barriers included phone access low digital literacy and unreliable electricity. Door-to-door visits visits by trusted community health workers remained crucial especially for those without little access. Participants recommended hybrid strategies including public announcements, community theater and visual media. To conclude, a hybrid communication approach that integrates digital tools with traditional outreach is critical in peri-urban Karachi. Interventions must prioritize equitable digital access and culturally relevant strategies to bridge health literacy gaps and improve maternal and child health outcomes.

## Introduction

Health literacy is an important social determinant of health that serves as the foundation for effective health promotion[1]. The World Health Organization (WHO) defines it as an individual’s ability to obtain, comprehend and apply health information to maintain and improve their own health while also contributing to the well-being of their families and communities[2]. Globally, health literacy has received substantial attention due to its direct impact on health outcomes[3]. According to a recent report from the US Department of Education and the National Centre for Education Statistics, one in four parents have poor health literacy, resulting in issues such as poor child nutrition, medication errors, and higher emergency department visits[4]. This challenge is more pronounced in Low- and Middle-Income Countries (LMICs), where inadequate education, high healthcare costs and various socio-cultural barriers further hinder access to reliable health information[5,6].

Health related information can be disseminated through various mediums including text messages, phone calls, television, newspaper articles, and community group gatherings[7]. Community-based approaches, particularly home-to-home visits by community health workers (CHWs), have proven to be highly effective in improving health literacy, particularly in limited-literacy settings[8]. A recent study among the rural residents of South Asia and Africa explored the effectiveness of participatory learning and action cycles, in which local facilitators made home visits to educate pregnant women about safe birth practices and newborn care; this resulted in a significant increase in knowledge, confidence, and the acceptability of recommended practices[9].

While these traditional outreach methods remain important, digital health messaging has emerged as a powerful complementary tool for health promotion[10]. A systematic review highlighted that social media plays a significant role in health communication by providing peer, social, and emotional support to the general public, while also facilitating the exchange of health messages[11]. Knop et al. further reported that Mobile Health (mHealth) interventions can improve antenatal care attendance and the timeliness of child immunizations in LMICs, reinforcing the potential of digital tools in maternal and child health[12]. However, a global scoping review of digital health interventions found that most evidence on digital health promotion is disproportionately focused on high-income countries[10]. This imbalance limits the applicability of the findings to LMICs, where differences in healthcare infrastructure and access to digital tools are also more pronounced[13]. As of January 2025, Pakistan Telecommunication Authority reports that mobile cellular subscriptions cover approximately 79.30% of the population, while mobile broadband penetration stands at 57.08%, highlighting a gap in high-speed internet access[14]. Kazi et al. also assessed mobile phone access in both urban and rural regions of Pakistan and found that while mobile phone ownership is high, smartphone penetration remains limited. This indicates that a significant portion of the population still relies on basic phones with limited functionality, posing challenges for digital health interventions[15].

In peri-urban communities of Karachi, where digital penetration is further limited, door-to-door outreach remains a vital method for health promotion. However, this method is often expensive, resource intensive, and requires significant time[8,16]. Given these challenges, it is imperative to explore the perceptions of women and community stakeholders regarding appropriate health communication channels in these areas. This can enhance health message dissemination and guide the development of targeted interventions to bridge healthcare knowledge gaps and improve maternal and neonatal outcomes in the future.

## Materials and methods

### Study design and setting

This exploratory qualitative study was conducted from August to December 2024 in four peri-urban sites situated along the coastal belt of Bin Qasim Town, Karachi. Collectively, these sites have a population of approximately 550,000 people, predominantly of Sindhi ethnicity[17].

For most residents, fishing serves as the primary source of income. These areas also have Primary Health Centres (PHCs), which are a part of maternal and child surveillance run by the Department of Paediatrics and Child Health at the Aga Khan University since 2010. As a part of this program, CHWs from the same communities conduct quarterly household visits to document vital events such as pregnancies, births and deaths. They also provide the local residents with preventive and promotive messages about antenatal care, skilled delivery, postnatal care, nutrition and childhood immunization. All data is collected electronically using an Android-based application, which is then securely stored in a cloud database.

### Study Participants

This study included two participant groups: women and community stakeholders.

A system-generated list of married women of reproductive age was extracted from the surveillance database. Only those who had resided in one of these catchment areas for at least six months, had at least one child under five years, and were able to communicate in Urdu, were eligible for inclusion. From the final list, 20 women were randomly selected, and then the first 10 eligible participants were approached. If a woman declined, the next eligible individual on the list was contacted.

Community stakeholders, including political leaders, teachers, social activists, and religious figures, were identified using purposive sampling. Only those with a minimum of one year of community service in the catchment area were eligible for inclusion. Their identities and credentials were verified by personnel at the PHCs.

### Data Collection

Data for this study was collected through focus group discussions (FGDs) with women and Key Informant Interviews (KIIs) with community stakeholders. The FGD and KII guides were developed after an extensive review of relevant literature from LMICs (Supplementary File 1 and 2).

After receiving approval from the Ethics Review Committee of Aga Khan University (AKU) (ERC # 2024-10183-29874), the eligible women and community stakeholders were invited on a pre-decided date, and time at the PHC located at each site. The data was collected from 21^st^ August 2024 to 23^rd^ December, 2024. Informed consent was obtained for both participation in the discussion and audio recording. In particular, participants without formal education were requested for thumb impressions as a means of consent, and an educated witness also signed their consent forms. During these interviews, women were asked about their sources of health information, preferred mediums of communication, and the facilitators and barriers they encountered in accessing health messages. We also explored their perspectives on the role of community leaders in disseminating health-related messages. Community stakeholders were questioned about the effective methods of communication, their roles in circulating health messages, the challenges they faced, and their recommendations to improve outreach.

The duration of each interview and discussion was 30 to 45 minutes. Two researchers were always present at the time of interviews to ensure rigor. One researcher conducted the interview, while the other took detailed notes. SK and HM fully transcribed the audio recordings into Romanized Urdu to preserve the original meaning of the transcripts. The transcripts were subsequently translated into English by SK and HM for coding and inclusion in the manuscript. All translations were reviewed by SK, AK and HM to ensure accuracy and minimize potential misinterpretations.

The study team aimed to conduct 1-2 FGDs per site with 7-8 women per FGD and 1-2 KIIs per site, or until data saturation was achieved in these communities. Saturation was achieved by the seventh FGD and KII, as confirmed through a review of the audio recordings, detailed notes and interview transcripts by the study team. This flexible approach also justified the final sample size.

### Data Analysis

The six-step method of thematic analysis described by Braun and Clarke guided the analytical process[18]. This method involved familiarizing with the data, generating codes, clustering codes into themes, reviewing and refining the themes, assessing their significance, and finally reporting the findings. Interview transcripts, notes and observations were thematically analysed using an inductive approach by SK, HM and AK. To facilitate concurrent data collection and analysis, transcripts were transcribed and translated into English by SK and HM after each interview and discussion respectively. The coding process was carried out using Dedoose software version 9.2.22 by SK, AK and HM. The data was systematically structured and categorized into themes that aligned with the research objectives, including the growing but constrained role of digital platforms, various facilitators and barriers to effective health message dissemination and community-suggested solutions. The researchers (SK, AK and HM) iteratively refined the codes and identified additional themes as the analysis progressed. This process involved multiple coders revising the codebook through team consensus, ensuring consistency in coding across all transcripts. To maintain qualitative rigor, transcripts were reviewed multiple times by SK, AK and HM and any discrepancies were resolved collaboratively. After a comprehensive review, a final set of codes was developed, accompanied by relevant data extracts that were organized into broader categories using MS Excel. At the end, senior authors (ZH, IN and FJ) reviewed these themes to ensure accuracy and validity.

### Positionality Statement

As a qualitative exploratory study, careful attention was given to issues of identity, positionality, and reflexivity. The research team comprised six individuals with diverse levels of experience in maternal and child health. Interviews were conducted by early-career researchers with medical backgrounds (SK and HM) under the guidance of senior researchers with public health expertise (AK, ZH, IH and FJ), ensuring both ethical integrity and methodological rigor.

Researcher positioning—often conceptualized in terms of being an “insider” or “outsider” to the study population—was complex in this context. Although the researchers and participants were from the same city, the researchers’ relative material privilege, largely shaped by their class position, created a disconnect from the lived experiences of the marginalized and vulnerable communities under study. Their understanding of the topic was primarily shaped by professional exposure rather than personal experience.

The team’s affiliation with Aga Khan University and, by extension, the PHC, also played a significant role in shaping the research process. This institutional connection fostered trust and facilitated community access, owing to the university’s established presence in the area. However, it also highlighted differences in social, economic, and professional status between the researchers and participants. To mitigate the influence of a predominantly biomedical perspective, deliberate efforts were made to value participants’ voices. Interviews were conducted in a way that recognized participants as experts in their own lives, encouraging open and honest sharing of their experiences and perspectives.

## Results

We approached 220 women of which 55 agreed to participate in the study. We conducted 7 FGDs with women who consented and 7 KIIs with community stakeholders. Table 1 summarizes the demographic information of the 55 women and 7 stakeholders included in the study. The mean age of women was 27.05 years (SD=5.43), with ages ranging from 19 to 40 years. About 43.6% (n=24) of the women had no formal education and nearly half of the women (n=25) owned mobile phones. Among mobile phone owners, 15 (60%) were smartphones users. Of these, 11 (73.3%) had attended some formal schooling while 4 (26.7%) had no formal education.

**Table 1:** Baseline characteristics of the participants enrolled in the study.

| **Women (n=55)** | **n(%)** |
| --- | --- |
| **Age in years (Mean±S.D<sup>a</sup>)** | 27.05±5.43 |
| **No. of children under 5 years** |  |
| 1 | 41 (74.5%) |
| 2 | 10 (18.2%) |
| 3 | 4 (7.3%) |
| **Education** |  |
| No formal Education | 24 (43.6%) |
| Primary | 10 (18.2%) |
| Secondary or higher | 21 (38.2%) |
| **Owner of mobile phones** | 25 (45.5%) |
| Basic mobile phone | 10 (40%) |
| Smartphone | 15 (60%) |
| **Community Stakeholders (n=7)** | **n(%)** |
| **Age, years; median (range)** | 27 (22-60) |
| **Gender** |  |
| Male | 3 (42.9%) |
| Female | 4 (57.1%) |
| **Education** |  |
| Secondary | 1 (14.3%) |
| Higher Secondary | 6 (85.7%) |
| **Occupation** |  |
| Social Activists | 4 (57.1%) |
| Vaccinator | 2 (28.6%) |
| Teacher | 1 (14.3%) |
| **Community work, years; median (range)** | 11 (5-40) |

a S.D. = Standard Deviation

The community stakeholders had a median age of 27 years (range 22-60), and most of them had completed higher secondary education (n=6, 85.7%). The various occupations among stakeholders included social activists (n=4, 57.1%), vaccinators (n=2, 28.6%), and teachers (n=1,14.3%). They also had extensive experience of working within their community, with a median duration of 11 years (range 5-40 years).

Thematic analysis revealed three major themes from both the participant groups including (i) the growing but constrained role of digital platforms, (ii) facilitators and barriers of effective health message dissemination and (iii) community-suggested solutions.

### The growing but constrained role of Digital Platforms

Participants had varying perspectives on the use of digital platforms for accessing health-related information. While some women mentioned barriers to accessibility of mobile phones, smartphone users highlighted smartphones as one of the main digital platforms for acquiring health related information as well as for receiving calls from the PHCs. One participant stated,

> *I use it to consult a doctor regarding my child*.*”* (Woman #2 in FGD 5).

Smartphone users, in addition, reported frequent use of social media applications such as YouTube, Facebook and WhatsApp. A few community leaders also expressed the use of WhatsApp groups for sharing health tips and addressing vaccine hesitancy in the communities. A key informant described,

> “*We have our own WhatsApp group, and we post statuses there. My friends are part of the group, and some of them (group members) are even (vaccine) refusers. When they see these messages, they sometimes agree with us*.*” (KI6)*.

Other social media platforms were also acknowledged by women and community stakeholders. In particular, a social activist stated that NGO workers and community members have been creating vlogs to showcase local skills and address health topics,

> *“Now we have worked on introducing our educated youth to YouTube and Facebook, where they (NGO workers) create vlogs related to this community and their skills*.*” (KI1)*

Beyond mobile-based communication, participants also suggested integrating health-related information into news segments on private TV channels during peak viewing times. A respondent stated,

> “*Many people watch the news. It would be ideal to have health-related information on the same channel. Husbands turn on the TV in the morning before going to work, so many people would see it. In the evening, news is also watched…especially around 7 or 8 p*.*m*.*…The 9 p*.*m. news is also watched by many*.*” (Woman #8 in FGD 2)*.

### Facilitators of Effective Health Message Dissemination

The use of visual content, such as videos and posters, was frequently mentioned as an effective way to convey health messages in these communities. Women particularly emphasized the need to adapt this content to the linguistic diversity of the locals for better understanding. A participant explained,

> “*Watching videos isn’t difficult; if it’s just written, where’s the enjoyment? Watching is still enjoyable. We can listen to the video while doing work, which makes it easier. Reading requires taking out time*.*” (Woman #5 in FGD 1)*.

One stakeholder stated,

> *“You put up posters, and whoever sees them will know, for example, that this vaccine is good and not harmful. Gradually, they (the community) become convinced*.*” (KI5)*

Furthermore, both women and stakeholders identified that the established trust between CHWs and the local community influences the reach of health messages. While stakeholders mentioned the strong relationships they have built within the community, local women acknowledged the reliability and trust in local health workers. One participant explained,

> *“We listen to them (health workers—polio workers, Sindh workers, and CHWs). They come regardless of time and explain things to us, which benefits us*.*” (Woman #8 in FGD 2)*.

### Barriers of Effective Health Message Dissemination

Limited access to mobile phones was mentioned as a significant barrier to health promotion by many women in the discussions. One woman explained,

> *“We don’t have phones, so videos on WhatsApp don’t make sense to us. How would we watch them?” (Woman #5 in FGD 7)*.

On the other hand, smartphone users and the stakeholders highlighted power supply issues, as an important barrier to access information provided through phones or television. Additionally, women expressed concerns about their limited time for using mobile phones, as household responsibilities often took priority.

> *“We don’t get much time to use them (TV and mobile phone) … We only watch TV or use mobile phones when we have time, and sometimes we don’t have time for days*.*” (Woman #5 in FGD 3)*

Cultural norms, gender dynamics and limited education also hinder healthcare decision-making. Stakeholders highlighted how men’s opinions often outweigh women’s in household decisions, restricting women’s autonomy despite being aware of the health benefits heard through various communication channels. A vaccinator described this challenge,

> *“Men’s opinions hold more weight. For example, when we go for vaccinations, women often say, ‘Ask my husband first, then I’ll proceed*.*” (KI3)*

### Community-suggested Solutions

Given the limited access to modern communication tools and the barriers that hindered their use, door-to-door campaigns have remained crucial for disseminating health messages. Since traditional lifestyles still prevail strongly in the community and not every household has a mobile phone, one participant emphasized the following:

> *“A team should come to our homes and share information. Many households have no knowledge of these things. So, if a team visits, they can explain things properly and get our attention by saying, ‘Look, if you focus on this and do it, things will improve for you*.*” (Woman #8 in FGD 3)*.

Stakeholders also highlighted the unique benefits of face-to-face interactions in building rapport with the community, as a vaccination supervisor explained,

> “*The understanding we can achieve through face-to-face interactions can’t be replicated through mobile phones. Face-to-face, we can understand their mindset, and they can understand ours*.*” (KI6)*

While face-to-face interactions were widely preferred, some participants acknowledged the potential of digital platforms for health message dissemination. A user suggested creating WhatsApp groups for sharing health messages, similar to how she had one for her children’s school, where school notices were shared.

Beyond direct communication, women also acknowledged the role of public announcements in community spaces for conveying health-related messages. As one woman explained,

> *“If there’s a vaccination team visiting, the announcement is made in the mosque. Many people hear it and go there*.*” (Woman #1 in FGD 6)*.

Lastly, women and stakeholders identified health camps and theatre performances as effective methods for directly delivering health messages and services to the community. A social mobilizer shared how a theatre performance was used to reach those who struggled to understand traditional messages, explaining,

> *“For communities that didn’t understand anything, we did a theatre performance…. It was easier, and it’s a simple way to make people understand. They all watch it with interest, and they understand it as well*.*” (KI5)*

## Discussion

The study explored the different perspectives of women and community stakeholders on maternal and child health communication in peri-urban Karachi. Findings highlight that while smartphone users accessed health information through social media, many faced barriers like limited phone access, power supply shortages and time constraints in using digital media.

Additionally, household responsibilities often took priority, further limiting women’s ability to engage with digital platforms. Stakeholders similarly acknowledged the promise of digital tools but emphasized the continued need for trusted face-to-face interactions led by CHWs. These findings suggest that a hybrid approach, integrating digital strategies with existing door-to-door outreach led by CHWs, may improve health dissemination in similar settings. Our study reported an increasing use of digital platforms by smartphone users, particularly social media on mobile phones. Consistent with other LMIC studies including Pakistan[8,19], smartphone users in our study accessed information via WhatsApp, Facebook, and YouTube supporting the growing role of social media in health communication. Abrejo et al. further emphasized that digital interventions can help enhance decision-making, reduce dependency on CHWs, and lower travel costs[19]. However, trust in digital health information remains a concern; as shown in India where only 35% of users trusted social media content, reinforcing that digital outreach alone is insufficient[20].

While digital media holds significant potential for improving maternal and child health communication, limited mobile phone ownership and access remain major barriers[21]. Nearly half of the women in the current study did not own a mobile phone, and even among those who did, factors such as electricity shortages, digital literacy, and gender-based restrictions hindered their ability to engage with online health content. These findings mirror research from India’s BIMARU states and rural Nepal, where mHealth interventions have shown promise in improving maternal and child health outcomes but are impeded by socio-economic, infrastructural, and cultural disparities such as poor internet connectivity, lack of electricity and geographic constraints[22,23]. Innovative solutions have been introduced to address these challenges; for instance, in rural India and Bangladesh, solar-powered internet solutions have been implemented to ensure reliable connectivity in areas with frequent power outages[23].

Additionally, community stakeholders emphasized their pivotal role in health communication, mentioning trust and familiarity as key factors in effective message dissemination. This is similar to a recent study which reported that stakeholder engagement in health communication enhances the trustworthiness and acceptance of health messages[24]. A study conducted in rural districts of Pakistan also found that local influencers, particularly religious leaders, can add credibility to health messages, while CHWs aware of the local cultural practices can effectively address vaccine hesitancy[25]. Perspectives from SAARC countries further report the important role of CHWs in improving maternal and neonatal healthcare outcomes, as their involvement in antenatal and postnatal care has significantly improved birth outcomes and newborn care practices in the underserved communities[26,27].

Previous research has emphasized the effectiveness of traditional communication methods in disseminating health information, particularly in communities with limited access to digital technologies. A recent qualitative assessment of the Sukh Initiative in Karachi, Pakistan highlighted the effectiveness of door-to-door services and free camps in promoting contraceptive use among married men and women[28]. The current study has similar findings where door-to-door campaigns remain a vital approach for disseminating health messages. Furthermore, public announcements in community spaces play a pivotal role in public health promotion, as expressed by the study participants. A scoping review also identified various health-promoting interventions conducted within mosques, emphasizing their effectiveness in fostering positive health behaviours and addressing communication barriers within Muslim communities[29].

Various organizations such as WHO and CDC suggests the use of interactive strategies to enhance understanding and engagement for health communication and behaviour change interventions[30,31]. Participatory communication approaches, which involve engaging communities through interpersonal interactions, using visual content and multi-channel media, have been shown to cultivate community interest and active participation in health-related matters[32]. A rapid review on participatory health interventions across various public health domains emphasized the benefits of integrating digital formats, demonstrating their role in fostering greater community engagement[33]. Additionally, participatory communication strategies like community theatre, particularly in young adults, have proven to be effective in promoting well-being and raising awareness of critical healthcare issues[34]. Our findings resonate with these studies, reinforcing the continued relevance of traditional communication methods and emphasizing the need for hybrid strategies that integrate both conventional and digital approaches to enhance health message dissemination and reach.

A major strength of our study is the inclusion of both message recipients and disseminators, offering a comprehensive overview of communication dynamics. Regular and detailed debriefings during data collection also enhanced reflexivity and analytical rigour. However, since our focus was in maternal and child health messages, we included only women with at least one child under five; this targeted inclusion may not fully capture the preferences of older women. Additionally, our study was conducted in a specific region of Pakistan, potentially limiting the generalizability of findings to other areas or countries with different socio-cultural contexts.

## Conclusion

In conclusion, while digital platforms present opportunities for expanding maternal and child health communication, they must complement, not replace face-to-face methods. A hybrid communications strategy grounded in local realities and community trust is crucial in building health literacy gaps and improving maternal and child health outcomes in underserved areas.

## Data Availability

The datasets used during the current study are available from the corresponding author on reasonable request.

## Acknowledgements

We are thankful to all frontline health workers for their support in conducting the interviews and discussions at the community PHCs.

## Supporting Information

Supplementary File 1: This contains the guide used for FGDs conducted with women.

Supplementary File 2: This contains the guide used for KIIs conducted with community stakeholders.

